# The experiences of 33 national COVID-19 dashboard teams during the first year of the pandemic in the WHO European Region: a qualitative study

**DOI:** 10.1101/2021.11.23.21266747

**Authors:** Erica Barbazza, Damir Ivanković, Karapet Davtyan, Mircha Poldrugovac, Zhamin Yelgezekova, Claire Willmington, Bernardo Meza-Torres, Véronique L.L.C. Bos, Óscar Brito Fernandes, Alexandru Rotar, Sabina Nuti, Milena Vainieri, Fabrizio Carinci, Natasha Azzopardi-Muscat, Oliver Groene, David Novillo-Ortiz, Niek Klazinga, Dionne Kringos

## Abstract

**Background:** Governments across the WHO European Region prioritized dashboards for reporting COVID-19 data. The ubiquitous use of dashboards for public reporting is novel. This study explores the development of COVID-19 dashboards during the pandemic’s first year and common barriers, enablers and lessons from the experiences of teams responsible for their development.

**Methods:** Multiple methods were applied to identify and recruit COVID-19 dashboard teams using a purposive, quota sampling approach. Semi-structured group interviews were conducted between April– June 2021. Using elaborative coding and thematic analysis, descriptive and explanatory themes were derived from interview data. A validation workshop with study participants was held in June 2021.

**Results:** Eighty informants, representing 33 national COVID-19 dashboard teams across the WHO European Region participated. Most dashboards were launched swiftly in the first months of the pandemic, between February–May 2020. The urgency, intense workload, limited human resources, data and privacy constraints, and public scrutiny were common to the initial development stage. Themes related to barriers or enablers were identified pertaining to the pre-pandemic context, pandemic itself, people and processes, software, data, and users. Lessons emerged around the themes of simplicity, trust, partnership, software and data, and change.

**Conclusions:** COVID-19 dashboards were developed in a learning-by-doing approach. The experiences of teams signal initial under-preparedness was compensated by high-level political endorsement, the professionalism of teams, accelerated data improvements, and immediate support of commercial software solutions. To leverage the full potential of dashboards, investments are needed at team-, national- and pan-European-level.

## Introduction

Governments, as the stewards of healthcare systems, have the chief responsibility to protect and promote the health and well-being of the population [1]. The stewardship role includes collecting and reporting relevant information and supporting its use as (performance) intelligence by all healthcare system actors, including the general public [2,3]. This task takes on new pertinence in the context of a public health emergency such as the COVID-19 pandemic [4-7]. Governments worldwide prioritized tools for delivering pandemic-related information, most often through the development of public web-based dashboards [8,9].

Dashboards can be characterized as a dynamic, visual display of key performance indicators, arranged on a single screen for viewing at-a-glance [10-12]. The ubiquitous use of dashboards as a public reporting tool during a pandemic is novel [13,14]. Nonetheless, the use-case for dashboards in a pandemic is clear. Contrary to static reporting, their dynamic nature iterates content and its display daily, evolving with the pandemic’s stages [15]. By design, dashboards can manage large data sets, which together with their near–real-time reporting capabilities, is conducive to a pandemic’s information urgency [8]. And, when paired with geographic information systems (GIS) and interactive drill downs, dashboards are critical for local monitoring, reporting and decision-making [12].

In the health sector, dashboards have traditionally been used for internal purposes, assisting managers in strategic and operational decision-making, particularly in hospitals [16], and supporting clinicians in clinical care and quality improvement [11]. There are also notable examples of public web-based dashboards for international health system benchmarking [17-22]. In contrast to COVID-19 dashboards, these have traditionally not been updated in near–real-time. Previous studies have explored optimizing the design of dashboards in healthcare [10], their effects on quality in clinical practice [11], and their development and implementation cycles [15,23]. In the context of the COVID-19 pandemic, scientific accounts have documented the technical development of dashboards [9,24-28] and their applications in clinical practice [29,30]. From a healthcare performance intelligence perspective, our research group *HealthPros* [31], has conducted international comparative research on COVID-19 dashboards, exploring features common to highly actionable dashboards [32] and their evolution over time [33,34].

In the European region, several actors (e.g. World Health Organization (WHO), European Centre for Disease Control, and Eurostat) have led initiatives for multi-country COVID-19 surveillance, setting new methodological standards for data collection and comparative reporting (e.g., [6,35]). To-date, describing the processes of developing COVID-19 dashboards has predominately focused on individual country accounts, typically reported by the media (e.g., [36-39]). Studying the development process is critical to ensure that first-hand experiences of developing COVID-19 dashboard will not be lost. Several systematic approaches to capture pandemic experiences have already been published, offering important insights from the perspective of healthcare providers [40-42], patients [43,44], and the general public [45-47].

To support governments during the current pandemic and to better prepare for future health threats as well as other potential uses of dashboards, our research group set out to conduct a multi-country study on the process of developing COVID-19 dashboards across Europe and central Asia. To do so, we partnered with the WHO Regional Office for Europe, as a key convening actor in the region and counterpart of our target healthcare system stewards. With this aim, the study was guided by two research questions: (1) How can the development process of COVID-19 dashboards during the first year of the pandemic be described from the perspective of the teams responsible for development? And (2) what common barriers, enablers and lessons can be derived from their experiences?

## Methods

### Design

The study adheres to the Consolidated Criteria for Reporting Qualitative Research [48]. To retrospectively examine the development process, a series of semi-structured group interviews were undertaken with COVID-19 dashboard developer teams across the WHO European Region. We used multiple methods to identify and recruit dashboard teams using a purposive, quota sampling approach. Group interviews in the local language of teams, to the extent possible, provided rich, collective team reflections on experiences with the process [49]. To answer our research questions, we adapted an approach previously developed by the study team to describe and assess the actionability of COVID-19 dashboards [32]. We also drew on the findings of prior COVID-19 dashboard research to inform the characteristics/features explored [32] (Table 1).

**Table 1.** Overview of characteristics and features explored

| Focus by research question | Characteristics and features explored |
| --- | --- |
| Development process | Responsible organizations, teams and launch |
|  | Aims, users and content |
|  | Data sources and breakdowns (geographic, population) |
|  | Data display, interpretation and visualization |
|  | Future plans |
| Reflections on process | Barriers |
|  | Enablers |
|  | Lessons learned |

The study team included researchers from *HealthPros* and WHO European experts on health data. The multi-national nature of the study team ensured broad and complementary contextual, research, policy, and subject matter expertise. Team members conducting interviews (four women, six men) had previously researched COVID-19 dashboards [32,50,51]. They were trained in health services research and had prior research and professional experience in countries of the WHO European Region. Interviewers were collectively proficient in 13 languages used in the WHO European Region.

The research protocol was developed in accordance with the ethical requirements of the primary research affiliation to Amsterdam University Medical Centers of the University of Amsterdam. Participants provided written consent during the recruitment stage and verbally restated their consent at the start of their interview. Confidentiality was assured by assigning each participating dashboard team a random code (e.g., D1) and removing identifying information from verbatim quotes used throughout the paper.

### Sample of dashboards and informant recruitment

We defined our target sample of COVID-19 dashboard based on five criteria: (1) reporting of key performance indicators related to the pandemic; (2) use of some form of visualization; (3) publicly available in a web-based format; (4) reporting on the WHO European Region at national-level; and (5) development by a governmental organization or appointed authority. To increase the generalizability of our findings, we set out to recruit a geographically representative sample of COVID-19 dashboards from the region’s 53 Member States. We applied WHO’s European Health for All database [52] country subgroups and set a target of 50% representation from each: Members of the European Union (EU) before May 2004 (EU15) (n=15); Members of the EU after May 2004 (EU13) (n=13); the Commonwealth of Independent States (CIS) (n=12); and other European Region countries not included in these groups (n=13).

To identify COVID-19 dashboards, we consulted the affiliated country data sources of the international “WHO European Region COVID-19 Situation Dashboard” [53] and the sample from our previous COVID-19 dashboard work [32]. Additionally, we manually web-searched national, governmental COVID-19 webpages. Our target informants were referred to as “dashboard teams,” specified as members of core dashboard teams directly involved in developing and managing national COVID-19 dashboards, ideally from their inception.

Following identification of applicable dashboards, multiple methods were used to recruit dashboard teams for interviews. In case where contact details were listed, teams were reached directly via email and/or through social media (Twitter, Facebook, LinkedIn). Alternatively, where a direct contact was not publicly available, we applied a snowballing approach, soliciting the advice of existing networks on health information systems across Europe, including country focal points of the European Health Information Initiative (EHII) [54] and the Population Health Information Research Infrastructure (PHIRI) Project [55], among other experts known to the study team. To further the recruitment process, a public webinar, convening EHII and PHIRI focal points, was organized and hosted by WHO, in March 2021, during which the study protocol was presented. Approximately 45 participants attended and were promptly followed-up with bilaterally where applicable by the study team. For countries where these methods did not result in direct contact with dashboard teams, support from WHO Country Offices was obtained to contact their respective ministries of health, informing them of the study and soliciting their participation. In contacting all prospective informants, an overview of the study was provided in English or Russian (see Appendix 1). When possible, correspondence took place in the local language.

### Data collection

A detailed interview guide was developed by the first authors (EB,DI) and reviewed by the study team. Once finalized, a training session for interviewers was organised, with the aim to calibrate the interview process. Interviews consistently explored two main themes: 1) the dashboard’s development and 2) reflections on the process over the course of the first year of the pandemic (see Table 1). A brief version of the interview guide was prepared in English and Russian and provided to informants in advance of interviews (see Appendix 2). A structured pre-interview process was developed for interviewers to familiarize with the corresponding dashboard. For this, we adapted our descriptive COVID-19 dashboard assessment tool [32] and approach to scoring a dashboard’s actionability [56].

Between April and June 2021, 60-minute semi-structured group interviews were conducted with participating dashboard teams, either virtually or in-person. Dashboard teams were assigned to interviewers based on their language and context expertise. Interviews were conducted in pairs (a lead interviewer and second team member), where language competencies allowed for this and when the lead interviewer was not a first author (see Appendix 3 for the distribution of interviewers and languages used). With the agreement of informants, interviews were recorded, transcribed verbatim and, where applicable, translated to English. Transcripts were made available to informants upon request. Interview data were stored by the first authors. Fortnightly meetings were organized for interviewers to exchange impressions on the process and update on themes.

### Data analysis

The first authors analysed the translated interview transcripts to identify descriptive and explanatory themes using elaborative coding [57] and thematic analysis [58] in an Excel tool developed in the approach of Meyer and Avery [59]. The analysis process included familiarization with the data, development and piloting of a coding framework, independent coding, peer review, mapping and interpretation of results. The coding framework was aligned with the research questions and was developed based on the characteristics and features (Table 1) of the semi-structured interviews (level 1). Additional themes (level 2) were generated through open (unrestricted) coding. The first authors independently coded three test transcripts each, then collaboratively reconciled and revised their coding. The approach was reviewed and discussed with two team members (NK,DK) and an external qualitative researcher during the piloting phase. The transcripts were divided between the first authors for independent coding.

Once coding had been peer reviewed by the second coder, and reconciled by the first, a master dataset for analysis was developed. To analyse the dataset, the characteristics/features explored were equally divided among the first authors for re-reading, mapping and interpretation. In this process, we iteratively noted recurrent themes, as well as outliers. In reporting on the results by research question, verbatim quotes were extracted from the transcripts. To ensure validity of the findings, we employed different techniques, including researcher reflexivity, debriefing with all interviewers and reviews by the full study team. Additionally, validating findings with informants was organized through a virtual workshop, again hosted by WHO and attended by 55 study participations in June 2021.

## Results

### Sample of participating COVID-19 dashboards

Five WHO European Region Member States (5/53, 9%) did not have an applicable dashboard at the time of sampling. Three COVID-19 dashboards from non-Member States, yet representing territories within the WHO European Region, were identified through our sampling and included during the recruitment stage (Fig 1).

**Fig. 1.**
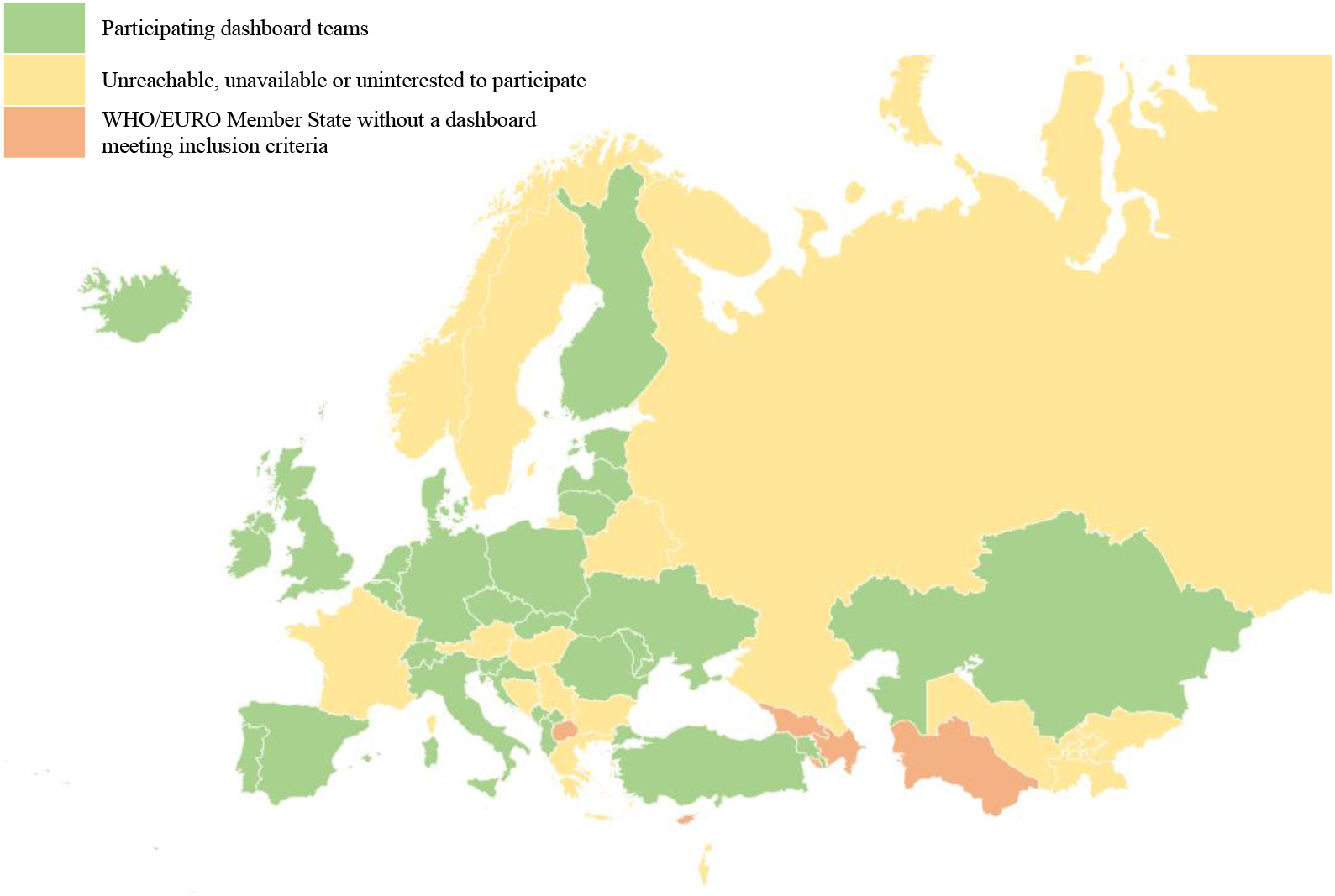
Geographic representation of participating COVID-19 dashboard teams. Note: Azerbaijan, Cyprus, Georgia, North Macedonia, and Turkmenistan did not have a dashboard meeting the inclusion criteria at the time of sampling. Non-WHO European Region Member States included in the sample Kosovo, Scotland and Liechtenstein. Liechtenstein is reported jointly on Switzerland’s dashboard.

In total, 33 dashboard teams participated in the study. See Appendix 3 for direct links to each dashboard. The dashboards represented 31 out of 53 WHO European Region Member States; 65% (33/48) of total Member States with eligible dashboards (Fig. 1). Seventeen Member States (17/48, 35%) were unreachable, unavailable or uninterested to participate. We met our target for representation in all but one regional subgroup, CIS countries, where a quarter of Member States did not have an applicable dashboard (3/12, 25%). Overall, participation rates of *eligible* dashboards by subgroups ranged 83% (10/12) of EU13 countries, 73% (11/15) of EU15 countries, 57% (8/14) of other countries/territories and 44% (4/9) of CIS countries.

Interviews engaged a total of 80 informants (45 men and 35 women). On average, two informants per dashboard were interviewed (range 1–8), with three or more informants contributing for 42% (14/33) of dashboard teams. Two dashboard teams provided written responses and two others required a second interview to finalize data collection. In one instance, an informant was directly involved in the development of two dashboards. Interviews were conducted in eleven languages (see Appendix 3).

### Description of the development process

#### Responsible organizations, teams and launch

The development of dashboards was predominately initiated by high-level officials, namely a country’s health minister, prime minister or president. Units of government or ministries of health (16/33, 48%), or national public health institutes (12/33, 36%) were most often appointed to lead their development (Table 2). In few instances, governments appointed other organizations (5/33, 15%) such as NGOs, private companies or academic institutions to develop the national dashboard, typically due to resource constraints. An awareness of the Johns Hopkins’ COVID-19 dashboard [60] and other national COVID-19 dashboards was often cited in connection with the initial request.

**Table 2.** Overview of development characteristics by participating dashboard team

| Country/<br>territory | Appointed<br>developing<br>organization<br>type | Launch date | Typology of content<sup>a</sup> |  |  |  | Open<br>Data<sup>c</sup> | Software |
| --- | --- | --- | --- | --- | --- | --- | --- | --- |
|  |  |  | Spread/death | Health<br>system | Vaccination | Other<sup>b</sup> |  |  |
| Albania | Gov/ministry | March 2020 | X | X |  |  | Yes | In-house solution |
| Andorra | Gov/ministry | October 2020 | X | X |  |  | No | In-house solution |
| Armenia | Other | May 2020 | X |  |  |  | No | In-house solution |
| Belgium | Public health | March 2020 | X | X | X |  | Yes | Google Data Studio |
| Croatia | Other | March 2020 | X | X |  |  | Yes | In-house solution |
| Czechia | Other | February 2020 | X | X | X |  | Yes | Google Data Studio |
| Denmark | Public health | April 2020 | X | X | X<sup>d</sup> |  | Yes | Esri ArcGIS |
| Estonia | Gov/ministry | March 2020 | X | X | X | X | Yes | Tableau |
| Finland | Public health | March 2020 | X |  | X<sup>d</sup> |  | Yes | Esri ArcGIS |
| Germany | Public health | March 2020 | X |  |  |  | Yes | Esri ArcGIS |
| Iceland | Gov/ministry | March 2020 | X | X | X |  | Yes | Infogram |
| Ireland | Gov/ministry | March 2020 | X | X | X |  | Yes | Esri ArcGIS |
| Italy | Gov/ministry | October 2020 | X | X |  |  | No | In-house solution |
| Kazakhstan | Public health | April 2020 | X |  |  |  | No | Qlik Sense |
| Kosovo | Public health | Early 2021 | X | X |  |  | No | Google Data Studio |
| Latvia | Public health | March 2020 | X | X |  |  | Yes | Infogram; Esri ArcGIS |
| Lithuania | Gov/ministry | November 2020 | X |  | X |  | Yes | Esri ArcGIS |
| Luxembourg | Gov/ministry | March 2020 | X | X | X |  | No | Qlik Sense |
| Malta | Gov/ministry | August 2020 | X |  | X |  | Yes | Infogram |
| Montenegro | Public health | March 2020 | X |  |  |  | No | Infogram |
| Netherlands | Gov/ministry | May 2020 | X | X | X | X | Yes | In-house solution |
| Poland | Gov/ministry | Early 2021 | X |  | X<sup>d</sup> |  | Yes | Esri ArcGIS |
| Portugal | Gov/ministry | March 2020 | X | X | X |  | No | Esri ArcGIS |
| Republic of Moldova | Gov/ministry | March 2020 | X |  |  |  | No | Esri ArcGIS |
| Romania | Other | March 2020 | X |  | X |  | Yes | In-house solution |
| Slovakia | Gov/ministry | March 2020 | X | X | X |  | Yes | In-house solution |
| Slovenia | Public health | Early 2021 |  |  | X |  | No | Microsoft Power BI |
| Spain | Public health | February 2020 | X |  | X |  | Yes | In-house solution |
| Switzerland | Other | November 2020 | X | X | X |  | Yes | In-house solution |
| Turkey | Gov/ministry | March 2020 | X | X |  |  | No | In-house solution |
| Ukraine | Gov/ministry | October 2020 | X | X | X |  | No | In-house solution |
| UK | Public health | March 2020 | X |  | X |  | Yes | In-house solution |
| UK–Scotland | Public health | April 2020 | X | X | X |  | Yes | Tableau |

Note: all data is according to the date of interview. Gov: government. UK: United Kingdom. <sup>a</sup>Refers to complete, anonymized datasets available directly from the dashboard, without additional permissions or requests. <sup>b</sup>Spread/death: cases, deaths, testing, reproduction rates, self-quarantine, etc. Health system: hospitalized, admitted to intensive care unit (ICU), hospital, ICU, ventilator capacity, personal protective equipment, etc. Vaccination: vaccination first and second dose. <sup>c</sup>Other: behavioural insights and social and economic impact. <sup>d</sup>Dedicated dashboard.

The launch dates of participating dashboards ranged three main time periods: February–May 2020 in the first months of the pandemic (24/33, 73%); late-2020 in parallel to the pandemic’s second wave across Europe (6/33, 18%); and early-2021 in connection with vaccinations (3/33, 9%) (Table 2). Dashboards that launched after the first months of the pandemic shared similar challenges, often data constraints or issues to identify a responsible organization. The unprecedented speed and workload involved in launching dashboards was a recurrent theme across teams. Many could vividly recount the initial days, recalling the level of uncertainty that characterized the process. As one informant described: “We were flying the plane as we were building it” (D31).

While some teams had prior experience with developing dashboards for internal use, most had never worked on dashboards intended for public reporting. Where possible, internal teams of data management units were re-purposed or new internal teams formed. Often, non-COVID activities were paused. For a quarter of dashboards (8/33, 25%), external teams were contracted to develop the dashboard. In two instances these teams were on a volunteer basis. Most teams started small, typically as one or two persons, though grew with time to about three to five core persons, and in some contexts, to more than twenty. The importance of multidisciplinary teams was emphasized, involving epidemiologists, public health specialists, information technology professionals, data analysts, policy experts and administrative staff. As teams expanded, additional expertise engaged included business intelligence and analytics experts, GIS specialists, user experience researchers and communication professionals. Support received from private front-end dashboard software suppliers was described as a critical addition to teams, especially in the early stages following their launch.

#### Aims, users and content

The dashboards were depicted as a vehicle for *informing*, but also as a tool for *partnering* with the public to “achieve greater participation of people in fighting the pandemic” (D9). Specific aims and target users were often implied rather than explicitly defined, with many citing the “chaotic” (D2) period that characterized the initial phase as a cause for this. Above all, the dashboards targeted the general public, though no dashboard team described having direct or formal contact with the public in the early development stage. Time constraints were consistently cited as the cause for this: “We would normally have done some user engagement and understand user needs, but the pace and expectation and demand to get the information out was so high” (D31). Other target users included national, regional and municipal officials, healthcare professionals, and the media.

In the early stages, following the dashboard’s launch, monitoring user analytics was not pursued for the common reason: “there wasn’t time for deep analysis of user behaviour” (D1). The intensity of the dashboard’s use was described to bring high expectations and “insatiable demands [for data]” (D8), new requests, and questions. While internal feedback mechanisms to dashboard teams were well-established, with dedicated pandemic crisis management teams or committees meeting daily, a structured process to manage the general public’s feedback was largely absent. Communication teams were described to play an important role in triaging these comments, predominately received via email and social media. However, the core dashboard team was typically tasked with providing technical replies; a demanding task given the magnitude and working pace.

Most dashboards (27/33, 82%) reported two or more types of content, most often spread/death and/or health systems or vaccination data (Table 2). More than half (21/33, 64%) had added vaccination datain late–2020 or early–2021, either as new tabs or separate dashboards. The latter was typically due to one of three reasons, or a combination thereof: a different organisation was mandated to coordinate and report on vaccinations; existing or new data collection infrastructure for vaccinations differed from the epidemiological system; and/or a different software solution was used. Beyond the addition of vaccination data, significant changes were avoided due to lack of time or concerns regarding the public’s reaction as they became accustomed to the dashboard and trusted the “original” version.

#### Data sources and breakdowns

“In the beginning, there were Excel spreadsheets” (D24). Many recounted similar intense manual data processing, especially countries/territories with more decentralized, less digitalized information systems. The availability, completeness, and quality of data ultimately played an important role in determining what indicators could be reported on, especially in the initial stages. Many described the trade-off between speed and quality, facing intense demands to publish data in near–real-time. This challenge intensified as the volume of data points increased with time. As one informant recounted: “Our data are usually ready for deployment 15–25 minutes before 4:00 pm, which is the time at which we usually deploy the data. So, we have that much time to curate 40 million records” (D10). Choosing to report open data was a political decision, typically made with view to ensure full transparency. More than half of the dashboards reported open data (21/33, 64%), meaning full data sets could be directly downloaded from the webpage (Table 2). Some went as far as to completely democratize their reporting: “The prime minister [country] sees the data at the same time as the guy down the road” (D10).

Data protection rules influenced a number of dashboard features including what indicators were reported on, data sources used, and geographic and population breakdowns applied. Clearances around what could be published were described to cause delays, as one dashboard team noted in reference to vaccination data: “We have had it ready for months, but right now the lawyers are debating, writing back and forth with the ministry” (D16). Fear of exposing personal health data was a repeat issue, with different interpretations of the lowest level of granularity when reporting cases locally, such as groups “larger than 20” (D16), “lower than 10” (D2) or “no less than 5” (D19).

Many described there was greatest interest to breakdown data to the local, municipal-level. This became increasingly relevant with the progression of the pandemic as infection control measures were introduced sub-nationally. The possibility to report data more locally also improved with time, as data collection processes became increasingly automated and of better quality. Persisting challenges included protecting privacy and ensuring smaller numbers were not misinterpreted by the public as low levels of risk. Resolving issues of incomplete denominators was also faced by some, describing outdated census data and challenges to record migrants, undocumented persons and seasonal workers. Beyond geographic breakdowns, age and sex disaggregation were common, though use of ethnicity and race-related, or socioeconomic status breakdowns were generally not pursued. This was in some instances due to lack of data, but more often, was a political decision. Specifically, some described uncertainty regarding its relevance for decision-making among the general public and fears of provoking discrimination.

#### Data display, interpretation and visualization

Dashboard teams relied on front-end dashboard display solutions developed either in-house (14/33, 42%) or commercially available (19/33, 58%). Most started with commercial solutions (21/33, 64%), typically Esri’s ArcGIS™ (11/33, 33%) which had a “COVID-dashboard module” by early-March 2020 (Table 2). Selecting a commercial solution was determined by a range of considerations: a team members’ previous experience with the software thus easing the learning curve; its availability for free (often for a limited period), meaning public procurement processes could be avoided; the proactive outreach and support of vendors; comparing the solutions used by other countries; and technical considerations, such as the degree of automation. Despite the speed-to-launch advantages of commercial solutions, they often faced limitations in terms of available templates and customisation. Most notably, the software selected was described to limit the range and types of visualisations and multi-language capabilities. Additionally, most commercial solutions were cloud-based, which was described by some as suboptimal, predominately due to data security concerns. For these reasons, four dashboard teams (12%) switched software with time.

Many described the task of visualizing data in a clear and understandable way as a challenge. Incorporating policy measures to explain data trends was seen by some as beyond their function of reporting *facts*. Providing detailed explanations and interpretations of the data were rather left to the media or what was described as “data enthusiasts” (D24) among the public. The dashboard was often part of a larger COVID-19 data and reporting ecosystem. Supplementary reporting efforts, mostly through static weekly situation reports, typically included additional indicators and more sophisticated analytics. These reports accommodated more text than the dashboard, making detailed explanations of data possible.

The dashboard teams described the importance put to preparing simple, easy to understand and interactive visualizations. In the early stages, visualizations were often not prioritized, as one developer described: “I can imagine maybe hundreds of other ways to visualize data describing the COVID situation. Unfortunately, because of lack of time, we decided to implement only the simple versions” (D1). Maps were consistently used to present local information, though privacy considerations also influenced visualizations, with some describing the challenge to avoid the suggestion that specific addresses were sites of cases and outbreaks.

#### Future plans

Discussing what is next for COVID-19 dashboards, four non-mutually exclusive scenarios were identified: (1) continuing to update existing dashboards, though less frequently with time; (2) further developing content (e.g., vaccines, wastewater studies), data management (e.g., automatization, quality, open data), design (e.g., visuals, organization) and user elements (e.g., low literacy levels, user behavior studies); (3) exploring non-COVID uses of dashboards for monitoring other communicable diseases (e.g., influenza) and registry data (e.g., cancers); and (4) preparedness planning, including investing in centralized data warehouses, in-house dashboard teams, coordinating across European countries and exploring alternative server and software options.

#### Barriers and enablers

Six main themes and fifteen subthemes were identified as recurrent barriers for some dashboard teams, yet enablers for others. These are briefly described below and listed in Table 3a–b.

**Table 3a.**
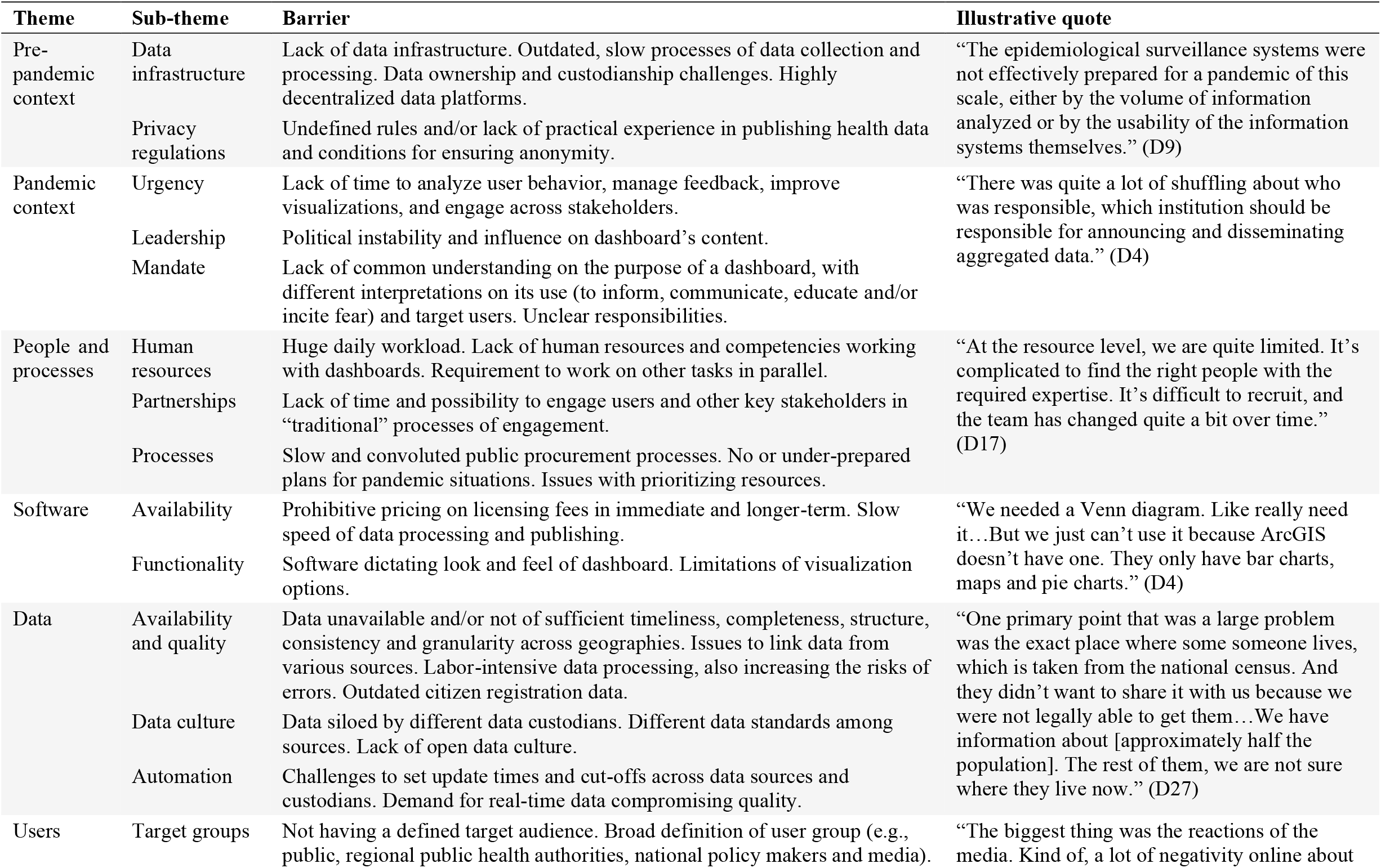
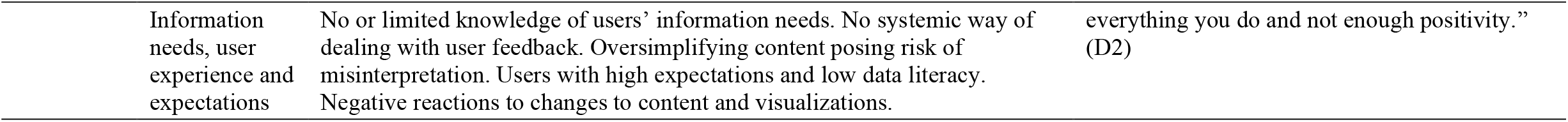
Summary of themes and illustrative quotes describing barriers

| Theme | Sub-theme | Barrier | Illustrative quote |
| --- | --- | --- | --- |
| Pre-pandemic context | Data infrastructure | Lack of data infrastructure. Outdated, slow processes of data collection and processing. Data ownership and custodianship challenges. Highly decentralized data platforms. | “The epidemiological surveillance systems were not effectively prepared for a pandemic of this scale, either by the volume of information analyzed or by the usability of the information systems themselves.” (D9) |
|  | Privacy regulations | Undefined rules and/or lack of practical experience in publishing health data and conditions for ensuring anonymity. |  |
| Pandemic context | Urgency | Lack of time to analyze user behavior, manage feedback, improve visualizations, and engage across stakeholders. | “There was quite a lot of shuffling about who was responsible, which institution should be responsible for announcing and disseminating aggregated data.” (D4) |
|  | Leadership | Political instability and influence on dashboard’s content. |  |
|  | Mandate | Lack of common understanding on the purpose of a dashboard, with different interpretations on its use (to inform, communicate, educate and/or incite fear) and target users. Unclear responsibilities. |  |
| People and processes | Human resources | Huge daily workload. Lack of human resources and competencies working with dashboards. Requirement to work on other tasks in parallel. | “At the resource level, we are quite limited. It’s complicated to find the right people with the required expertise. It’s difficult to recruit, and the team has changed quite a bit over time.” (D17) |
|  | Partnerships | Lack of time and possibility to engage users and other key stakeholders in “traditional” processes of engagement. |  |
|  | Processes | Slow and convoluted public procurement processes. No or under-prepared plans for pandemic situations. Issues with prioritizing resources. |  |
| Software | Availability | Prohibitive pricing on licensing fees in immediate and longer-term. Slow speed of data processing and publishing. | “We needed a Venn diagram. Like really need it...But we just can’t use it because ArcGIS doesn’t have one. They only have bar charts, maps and pie charts.” (D4) |
|  | Functionality | Software dictating look and feel of dashboard. Limitations of visualization options. |  |
| Data | Availability and quality | Data unavailable and/or not of sufficient timeliness, completeness, structure, consistency and granularity across geographies. Issues to link data from various sources. Labor-intensive data processing, also increasing the risks of errors. Outdated citizen registration data. | “One primary point that was a large problem was the exact place where some someone lives, which is taken from the national census. And they didn’t want to share it with us because we were not legally able to get them...We have information about [approximately half the population]. The rest of them, we are not sure where they live now.” (D27) |
|  | Data culture | Data siloed by different data custodians. Different data standards among sources. Lack of open data culture. |  |
|  | Automation | Challenges to set update times and cut-offs across data sources and custodians. Demand for real-time data compromising quality. |  |
| Users | Target groups | Not having a defined target audience. Broad definition of user group (e.g., public, regional public health authorities, national policy makers and media). | “The biggest thing was the reactions of the media. Kind of, a lot of negativity online about |
|  | Information needs, user experience and expectations | No or limited knowledge of users' information needs. No systemic way of dealing with user feedback. Oversimplifying content posing risk of misinterpretation. Users with high expectations and low data literacy. Negative reactions to changes to content and visualizations. | everything you do and not enough positivity.” (D2) |

**Table 3b.** Summary of themes and illustrative quotes describing enablers

| Theme | Sub-theme | Enablers | Illustrative quote |
| --- | --- | --- | --- |
| Pre-pandemic context | Data infrastructure | Electronic, centralized data flows (including hospital and laboratory data systems and infectious disease registries) with automatized data management. Ability to link data sources. | “The existing infrastructure, the central health information system and so on, those were definitely enabling factors. If we didn’t have them, or integration with the labs, with the hospitals, we would have been in the Stone Age.” (D24) |
|  | Privacy regulations | Supportive privacy and security legislation and practice. Enabling state of emergency conditions. |  |
| Pandemic context | Urgency | End-goal orientation contributing to committed and focused efforts. Not being perfectionist. “Once in a lifetime experience” as a motivator. | “[The] ‘chain of command’ was clear. That was an enabler, for sure. We all knew [the dashboard] was the official communication channel. It was practically considered and extended arm of the government.” (D15) |
|  | Leadership | Political and upper-management support and endorsement providing access to sufficient resources. High-level directives. |  |
|  | Mandate | Clear purpose and mandate. High degree of autonomy to dashboard teams. Easy access to data as well as to skills needed in-house and possibility to outsource for additional capacity. |  |
| People and processes | Human resources | Committed, competent, multi-disciplinary and proactive team. Prior experience with public reporting, dashboards and visualisations. | “This then led with an agile development to also change the relationships between people, who have become much less structured and hierarchical and have become much more intellectual and free.” (D12) |
|  | Partnerships | Improvements to intra- and inter-organisational communication and collaboration. Need-based stakeholder collaboration and engagement including communication specialists and decision-makers. |  |
|  | Processes | Flattened hierarchy. Streamlined and agile internal organization. Change of mentality towards a more operational one. |  |
| Software | Availability | Supportive technological solutions. Commercial software offered free-of-charge, for a period. Reusing existing solutions. | “There is no doubt that ten years ago the management of this pandemic would have been much more difficult, and now we have technology that has enabled us to fight the pandemic much more effectively.” (D9) |
|  | Functionality | Easy to build and automate. Flexible and easy to maintain once set up. Extensive vendor support. |  |
| Data | Availability and quality | Available, accurate and timely data of sufficient granularity. Ability to link data across sources and organizations. | “Because we are a data department...it’s very easy for us as we are data managers, all of us, and we have access to all the data. So, when we’re asked to do something, we don’t have to ask anybody. “Can you get me this data?” This access is very important for quick results.” (D16) |
|  | Data culture | Aligned data standards and methodologies. Culture of data interoperability, open data and secondary data use. |  |
|  | Automation | Streamlined data processes including automation of collection, processing and reporting. |  |
| Users | Target groups | Clearer definition of target audience. Separate dashboards or modules for different user groups with different information needs. Curious, rather than malicious, users. Partnership with media. Support and readiness for data-driven decision-making at all levels. Dissemination aids including social media platforms, high-level officials, transitional media, and data champions. | “We’ve had a good relationship with the media and there’s a short communication way from the public to us. So, if there’s something the public is insecure about or wants to know more about or wants [they can easily reach us], and that’s good.” (D26) |
|  | Information needs, user experience and expectations | Systemic approaches to researching user experience, implementing improvements and managing user feedback. Tradition of public using data for decision-making. |  |

#### Pre-pandemic context

The existing data infrastructure was a major challenge facing some dashboard teams, describing the limitations of traditional data collection and processing for near-real-time dashboard reporting. In contrast, teams working in settings with more advanced data systems and a culture of data (re)use, credited this as a contributor to their success. Similarly, some described the challenge resulting from highly decentralized data structures and processes, contributing to a lack of clarity around data ownership and custodianship. The limited level of preparedness for handling privacy regulations was a key barrier for some, whereas prior experience with interpreting privacy and security legislation in the context of public reporting, was an enabler for others.

#### Pandemic context

For most teams, chronic lack of time caused by the pandemic’s urgency and the demand to publish near–real-time data, meant analysing user behaviour, managing feedback, improving visualisations and engaging across stakeholders, became secondary. However, some teams did describe the constant urgency of the pandemic as a reason for streamlined processes and an end-goal orientation contributing to committed teams and focused efforts. The political context, including changes in leadership positions and the content demands of high-level decision-makers, was underscored as a barrier for some. So was the lack of a common understanding on the purpose and audience of dashboards and their position in the COVID-19 data ecosystem. Contrary to this experience, some teams credited high-level political endorsements for the development of national COVID-19 dashboards as a key enabler, providing ample resources and direction, yet autonomy, to the teams developing them.

#### People and processes

Seven-day workweeks, an immense workload and shortages of competent specialists hindered the development of dashboards in the experience of some teams. So did bureaucratic processes, including those for partnering with stakeholders, accessing and linking data, and engaging with public procurement. Conversely, having experienced data dashboards and business analytics teams in place, working across departments and organizations, and engaging new team members with necessary competencies were attributed as enabling factors in the experience of others. Flattening hierarchical structures and streamlining processes to facilitate decision-making, also were described to play a supporting role.

#### Software

Front-end dashboard software solutions occasionally impaired the development of dashboards, according to some teams, through limited visualisations, lack of multi-language functionality and other customization options as well as prohibitive pricing. For others, re-using existing in-house developed data analytics and dashboard tools (where available) provided more flexibility. When in-house options were not available, some teams described commercial software as an aid to accelerate the launch of dashboards. Waving initial fees and direct support from software vendors were cited as a key enabling factor at the outset.

#### Data

Publishing reliable, accurate, consistent and timely data proved challenging for many dashboard teams. Agreeing on data standards, including daily cut-off times, and the absence of granular data needed for reporting “close to home” and by different population sub-groups, were described as key hindrances. Conversely, the availability of (interoperable) data, coordination across data custodians, existing data (methodology) standards and cultures of secondary data use and open data publishing were described to benefit others.

#### Users

Managing users was described by some teams as a critical challenge, recounting intense scrutiny over the content of dashboards, issues of misinterpretation of data by users and negative reactions to mistakes as well as changes introduced. Some teams also detailed the high expectations of users (for real-time reporting), challenges in explaining methodologies (to lay audience) and lack of systemized processes to handle user feedback. For others, having clearly defined target audiences, knowing their information needs, engaging with media outlets, systematically improving user experience, and handling feedback was a way of partnering with users and overall, an advantage to the process. Transparency on methods and admitting mistakes, which inevitably happen at this volume and speed of work, were perceived as enablers.

#### Lessons learned

When asked to consider, with the benefit of hindsight, what dashboard teams would do differently or what advice they would offer others to best prepare for public reporting in the context of a crisis, five themes emerged. One theme was on the importance of simplicity, reporting only essential information, prioritizing content that can be easily interpreted and supported by explanations. A second was the importance of trust. Teams described the inevitably of errors, given the urgency and volume of data, and in effect, the importance of disclosing errors as they happen. Using open data and prioritizing data security and privacy were also important lessons for building user trust. Third, the necessity of working in partnerships, in agile and collaborative ways with system leaders but also across in-house units, other stakeholders and the target audience, was emphasized. A fourth theme was the importance of software and data. While intuitive and recognized at the outset, teams were continuously confronted with the parameters set by the software chosen and reminded of the importance to automate processes and invest in quality data. A last theme was about confronting the truly dynamic nature of dashboards, finding ways to learn from others to improve, adapt to the stages of the pandemic and embed dashboards within other reporting. Recurrent themes, lessons learned, and representative quotes are summarized in Table 4.

**Table 4.**
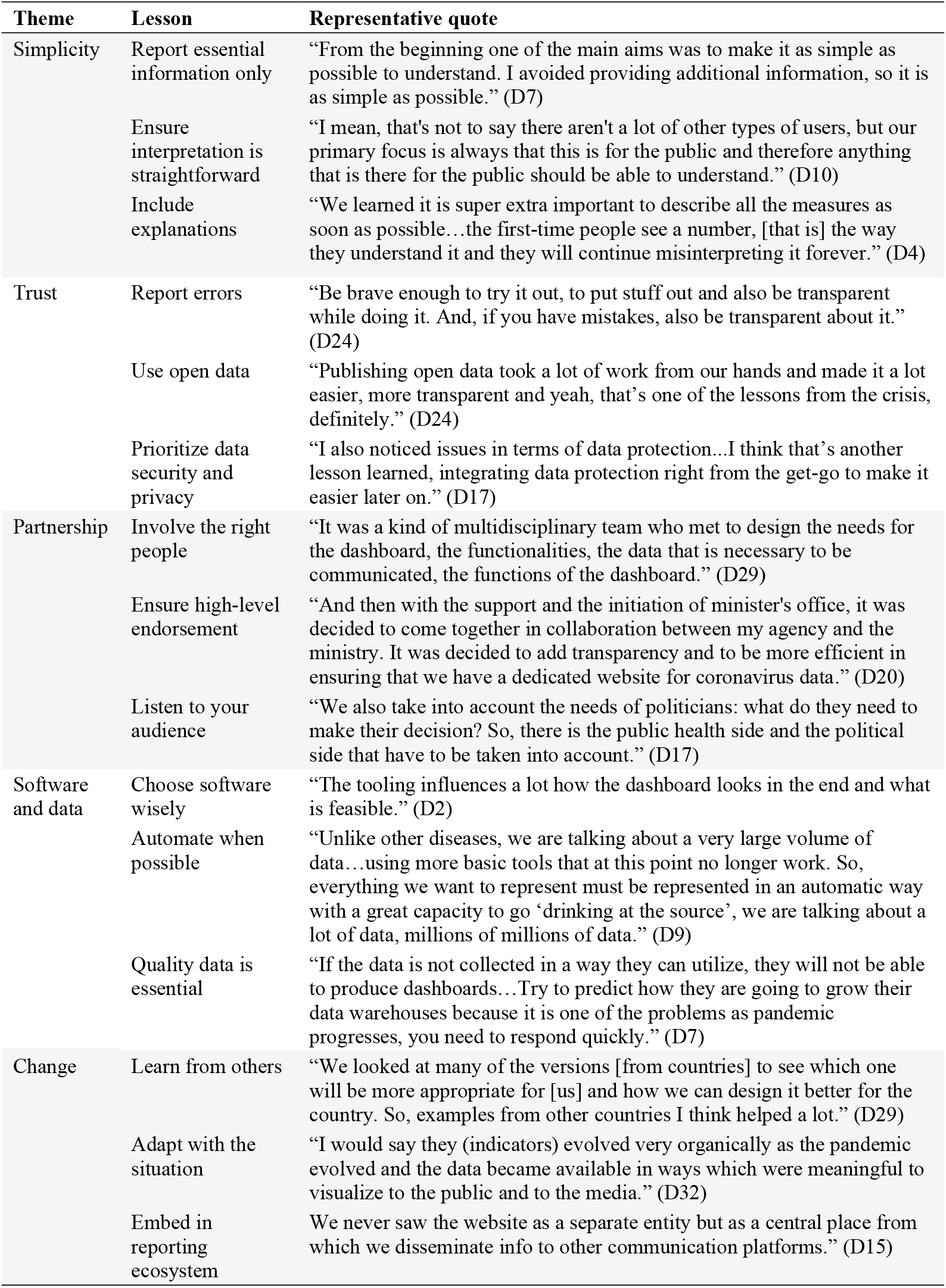
Summary of lessons learned, recurrent themes and representative quotes

## Discussion

In this study, we retrospectively explored the development of COVID-19 dashboards over the first year of the pandemic across the WHO European Region. The region itself is geographically vast, with countries/territories of wide ranging population sizes, health information systems at varied stages of development [61,62], diverse data and administrative cultures and traditions [4], among other key differences. Nonetheless, we found there were more similarities than differences in the development stories described by the 33 national COVID-19 dashboard teams we met with. The factors that hindered or facilitated the development process and resulting lessons learned also shared a number of commonalities.

The overall speed with which governments requested the development of public-facing COVID-19 reporting reflects the WHO European Region’s tradition of prioritizing good governance [63]. However, the ubiquitous use of dashboards for delivering COVID-19 data is found to be an international ripple effect, rather than the activation of pre-existing emergency response plans. The uptake of dashboards appears triggered by early adopters (e.g., [9]) and sustained by a proactive commercial software vendor market. As such, the statement of Boukaert et al. [4], “coping with the crisis has been first and foremost an issue of the national states, whereas the European voice has been weak,” holds true also with regards to COVID-19 public reporting tools, including dashboards. To stimulate pan-European collaboration, the convening role of international actors should be leveraged to advance a common approach to public reporting using dashboards and sharing of lessons across contexts.

Dashboards feed on relevant, quality data. In the literature, data sources are cited as an essential aspect for dashboard development [15,64,65]. Initially, national health information systems struggled to provide accurate, timely data for COVID-19 dashboards, a key challenge also reported by others [14,66,67]. Our findings signal the need for continued investment in national-level data sources that are integrated and interoperable, and digital infrastructure that spans secondary, primary and social care systems [14]. Preparedness to tackle data privacy and security issues, including practical applications of GDPR in the EU, needs further prioritization and should be guided by the aim to report data in a proportionate, ethical and privacy-preserving manner [14]. Cross-country efforts like the European Health Data Space [68] may serve to advance the development of common data standards, indicator sets, and methodologies to the betterment of national but also pan-European reporting.

While dashboards are an important communication tool, though like other digital technologies [14] they are not a silver bullet. Previous research has emphasized the features of dashboards must fit with its intended purposes [15]. The actionability of data for end users relies on the extent to which the information is communicated clearly and understood [69-71]. However, we observe differences in the extent to which teams explore, discuss and define dashboard’s purpose. Some saw it as presenting solely data (raw numbers) for the public to interpret themselves, versus others that endeavoured to provide explanations using narratives or visual methods. As reported in other studies, the way in which information is presented may change not only the subjective comprehension but also the objective understanding of the information [11,72,73]. Systematic approaches to explore user needs and use patterns are necessary, helping dashboards bridge the gap between being a managerial tool and a public reporting device.

The teams agreed that dashboards, and other near–real-time, web-based, interactive and visual reporting tools, are the likely future of public reporting. COVID-19 dashboards served as a proof-of-concept, including how much can be achieved with limited resources and high urgency. They have also served to flag imminent areas for improvement and new challenges like potentially harmful misinformation [74] Running dashboards in the longer-term and expanding their use into areas like resilience and recovery plans, and non-COVID monitoring, like on cancer, seasonal flu and patient safety, will have implementation and management costs which past studies have underscored as possibly prohibiting [15,16]. These were managed (or evaded) during the pandemic through mobilisation of emergency resources. However, the continued and expanded use of dashboards will require more intentional resource planning and investments.

### Study strengths and limitations

Working in partnership with WHO offered unique access to our target national governmental COVID-19 dashboard teams. The composition of the research team allowed for the use of an extensive range of languages during data collection, aiding recruitment and the richness of exchanges during interviews. The study captured COVID-19 dashboards at a critical point in their development, as teams were actively improving and making adjustments at the time of interviews. It means teams were still deeply immersed in the dashboards, making for little strain to recall processes of the prior year.

We acknowledge the following potential limitations. First, the size and composition of core dashboard teams varied across countries/territories and brought some variability to the profile and number of persons met with per dashboard, and ultimately, the nature of their experience. Second, group interviews stimulated joint reflections across teams, enriching data collection, though this approach could also induce group pressure resulting in socially desirable responses. Third, the findings are a snapshot of the first year of the COVID-19 pandemic and may not reflect the current state of implementation. Lastly, the study included national, government COVID-19 dashboards of the WHO European Region and may not be generalizable to the experiences of subnational dashboards, other types of developers, such as academia, independent initiatives, media outlets, or industry, and of other regions globally, in particular low-income countries.

## Conclusion

The study revealed more similarities than differences among the 33 participating COVID-19 dashboard teams from across the WHO European Region. A learning-by-doing approach described by the teams reflects the novelty of using dashboards as a tool for public reporting during a pandemic. The experiences of COVID-19 dashboard teams signal that initial under-preparedness was compensated by high-level political endorsement, teams’ own professionalism, accelerated data improvements and commercial software solutions. Recurrent barriers and enablers related to the pre-pandemic and pandemic context, people and processes, software, data, and users should inform investments at all levels—dashboard teams, national and pan-European–level. Lessons around the themes of simplicity, trust, partnership, software and data, and change indicate areas for taking action to fully realize a data-informed approach to stewardship using dashboards.

## Data Availability

All data relevant to the study are included in the article or uploaded as supplementary information. The data generated and/or analysed in the study are not publicly available due to participant anonymity.

## Declarations

### Ethical approval

Ethical requirements of the primary research affiliation to Amsterdam University Medical Centers of the University of Amsterdam, the Netherlands, for which an exception applies.

### Guarantor

EB, DI

### Contributorship

EB and DI contributed equally as co-first authors. EB, DI, KD, OG, DNO, NK, DK designed the study. All authors contributed to the tool development, sampling and/or recruitment of dashboards. EB, DI, OBF, MP, CW, BMT, VB, ZY, OG, NK conducted interviews, transcribed and translated interview data. EB, DI, NK, DK analyzed interview data. EB and DI prepared the manuscript. All authors reviewed drafts of the manuscript and approved the final version.

## Acknowledgements

The authors thank Jeanine Suurmond for methodological advice, and all the interviewees who generously shared their time to participate in the study: Assel Abakova; Anna Artsruni; Georgi Asatryan; Jonas Bačelis; Simon Bak; Senad Begić; Tania Boa; Lovro Bucić; Stefan Buttigieg; Alan Cahill; Fabrizio Carinci; Gianfranco Costabile; Phillip Couser; Igor Crnčić; Olgeta Dhono; Michaela Diercke; Mykola Dobysh; Marjolein Don; Anna Fumačová; Aram Ghulijanyan; Clare Griffiths; Veaceslav Gutu; Pouria Hadjibagheri; Luc Hagenaars; Scott Heald; Jonas Kähler; Kristjan Kolde; Martin Komenda; Kristina Kovačikova; Tanja Kustec; Terje Lasn; Sonia Leite; Renata Lenhardcziková; Pedro Licínio Pinto Leite; Jana Lepiksone; Mathias Leroy; Avet Manukyan; Elena Martinez; Graham McGowan; Emma McNair; Lorraine McNerney; Matej Mišik; Susana Monge; Teemu Möttönen; Maja Mrzel; Gints Muraševs; Hugo Agius Muscat; Martina Nagyová; Valentin Neevel; Aleksandar Obradović; André Peralta-Santos; Natalia Plugaru; Ane Radović; Anders Rasmussen; Raul Ritea; Leonardo Rocchi; Josep Romagosa; Giulio Siccardi; Fernando Simon; Margita Štāle; Alexandra Ștefănescu; Kristian Sufliarsky; Patrick Suter; Maríanna Þórðardóttir; Nataša Terzić; Vitaliy Trenkenshu; Alexander Ullrich; Zuzana Vallová; Jeroen van Leuken; Jana Vanagė; Giordano Veltro; Olivia Vereha; Solange Vogt; Liina Voutilainen; Martina Vrbiková; Marjana Vrh; Pauline White; Piotr Wlodarczyk; Kıvanç Yilmaz; Tatiana Zvonnikova.

---

**Consolidated Criteria for Reporting Qualitative Research (COREQ) Checklist**^**a**^

| # | Item | Question/description | Answer | Section |
| --- | --- | --- | --- | --- |
| **Domain 1: Research team and reflexivity** |  |  |  |  |
| **Personal Characteristics** |  |  |  |  |
| 1 | Interviewer/facilitator | Which author/s conducted the interview or focus group? | Team members conducted interviews: EB, DI, OBF, MP, CW, BMT, VB, ZY, OG, NK. | Methods |
| 2 | Credentials | What were the researcher's credentials? | EB, MSc*<br>DI, MD, MBA*<br>OBF, MSc MEd*<br>MP, MSc MD*<br>CW, MSc*<br>BMT, MSc, MD*<br>VB, MA*<br>ZY, MDP<br>OG, MSc, PhD<br>NK, MD, PhD<br>*were all PhD students | Methods |
| 3 | Occupation | What was their occupation at the time of the study? | All interviewers were members of an existing international network of healthcare performance intelligence professionals. Interviewers included researchers, doctoral students in their final year of training and principal investigators of the training network. | Methods |
| 4 | Gender | Was the researcher male or female? | Four women<br>Six men | Methods |
| 5 | Experience and training | What experience or training did the researcher have? | All interviewers had training in health services research, COVID-19 dashboard assessments and prior professional experience in WHO European Region countries. | Methods |
| **Relationship with participants** |  |  |  |  |
| 6 | Relationship established | Was a relationship established prior to study commencement? | Informants were contacted via email prior to interviews in the local language as far as possible. All were provided a study overview. A virtual information session (webinar) was organized to inform on the study and address questions of prospective participants. | Methods |
| 7 | Participant knowledge of the interviewer | What did the participants know about the researcher? e.g. personal goals, reasons for doing the research. | Informants all received a written overview of the study aims and objectives (Appendix 1) in English or Russian. The reasons for conducting the research were restated at the start of all interviews. | Methods |
| 8 | Interviewer characteristics | What characteristics were reported about the interviewer/facilitator? e.g. Bias, assumptions, reasons and interests in the research topic | At the outset of interviews, each interviewer present introduced themselves by their areas of research and affiliations. | Methods |
| **Domain 2: study design** |  |  |  |  |
| **Theoretical framework** |  |  |  |  |
| 9 | Methodological orientation and Theory | What methodological orientation was stated to underpin the study? e.g. grounded theory, discourse analysis, ethnography, phenomenology, content analysis | Characteristics/features explored were derived from an approach to describe and assess the actionability of COVID-19 dashboards previously identified by the study team and reported in detail elsewhere. Elaborative coding and thematic analysis were applied to analyze data. | Methods |
| **Participant selection** |  |  |  |  |
| 10 | Sampling | How were participants selected? e.g. purposive, convenience, consecutive, snowball | Purposive sampling according to 5 inclusion criteria | Methods |
| 11 | Method of approach | How were participants approached? e.g. face-to-face, telephone, mail, email | Multiple methods via email including direct contact, advice of experts (existing networks), snowballing of recommendations, WHO Country Office support. | Methods |
| 12 | Sample size | How many participants were in the study? | 76 informants<br>33 COVID-19 dashboards | Results |
| 13 | Non-participation | How many people refused to participate or dropped out? Reasons? | 17 target countries were unreachable, unavailable or uninterested to participate. | Results |
| **Setting** |  |  |  |  |
| 14 | Setting of data collection | Where was the data collected? e.g. home, clinic, workplace | Online via Zoom or Teams<br>One in-person interview | Findings |
| 15 | Presence of non-participants | Was anyone else present besides the participants and researchers | No | Methods |
| 16 | Description of sample | What are the important characteristics of the | Geographic mix, type of organization and number of team members are reported in | Results |
|  |  | sample? e.g. demographic data, date | Appendix 3 together with direct links to the dashboard discussed. |  |
| **Data collection** |  |  |  |  |
| 17 | Interview guide | Were questions, prompts, and guides provided by the authors? Was it pilot tested? | A detailed (scripted) interview guide was prepared for interviews. A training workshop was organized. | Methods |
| 18 | Repeat interviews | Were repeat interviews carried out? If yes, how many | Yes, two. | Methods |
| 19 | Audio/visual recording | Did the research use audio or visual recording to collect the data? | Interviews were audiorecorded with the agreement of participants. | Methods |
| 20 | Field notes | Were field notes made during and/or after the interview or focus group? | No. All interviews were transcribed verbatim to follow and translated to English as needed. | Methods |
| 21 | Duration | What was the duration of the interviews or focus groups? | 60 minutes. | Methods |
| 22 | Data saturation | Was data saturation discussed? | Regular team meetings of interviewers were organized to discuss progress in connecting with developers and recurrent themes. | Methods |
| 23 | Transcripts returned | Were transcripts returned to participants for comment and/or correction? | No. Transcripts were provided when requested by participants for their recordkeeping purposes. | Methods |
| **Domain 3: analysis and findings** |  |  |  |  |
| **Data analysis** |  |  |  |  |
| 24 | Number of data coders | How many data coders coded the data? | Two (EB, DI). | Methods |
| 25 | Description of the coding tree | Did authors provide a description of the coding tree? | Yes, 1 (Table 1) and 2 level coding was applied. | Methods |
| 26 | Derivation of themes | Were themes identified in advance or derived from the data? | Level-1 codes drew from characteristics/features explored in interviews (Table 1). | Methods |
| 27 | Software | What software, if applicable, was used to manage the data? | An Excel tool was developed. | Methods |
| 28 | Participant checking | Did participants provide feedback on the findings | Preliminary results were presented at a workshop in June 2021 attended by 55 informants. | Methods |
| **Reporting** |  |  |  |  |
| 29 | Quotations presented | Were participant quotations presented to | Confidentiality has been assured by assigning each participating | Results |
|  |  | illustrate the themes/findings? Was each quotation identified? e.g. participant number | dashboard developer team a random code D# (ranging 1 through to 33) and removing identifying information from the transcripts. |  |
| 30 | Data and findings consistent | Was there consistency between the data presented and the findings? | Findings are presented in the approach of the research questions and features/characteristics investigated (table 1) | Results |
| 31 | Clarity of major themes | Were major themes clearly presented in the findings? | Tables are used to present major themes (Table 2, 3, 4). Subheadings are used to improve clarity. | Results |
| 32 | Clarity of minor themes | Is there a description of diverse cases or discussion of minor themes? | Tables are used to supplement in-text descriptions of sub-themes (e.g., Table 3 and 4) | Results |

<sup>a</sup> Tong A, Sainsbury P, Craig J. Consolidated criteria for reporting qualitative research (COREQ): a 32-item checklist for interviews and focus groups. *Int J Qual Health Care* 2007;19:349–57.

## Appendix 1: Study overview

**Pandemic public reporting that is fit for purpose and use: A qualitative study of COVID-19 dashboards in the WHO European Region from the perspective of their developers**

### Background

Public reporting in the context of a pandemic is a core government function and critical to foster accountability, trust and transparency, and to support individuals to make informed, risk-minimizing behaviour changes. Since the outbreak of COVID-19, activity has surged worldwide to develop dashboards as dynamic, visual tools for communicating COVID-19 data. However, without careful selection of indicators and data collection, analysis and visualization, dashboards have the potential to mislead, misinform, and incite panic, or simply to be ignored.

In the first half of 2020, our international research network of Healthcare Performance Intelligence Professionals (HealthPros)^1^, launched a global study of COVID-19 dashboards. The study assessed 158 dashboards from 53 countries in July 2020. It also explored what makes a dashboard actionable, where actionability refers to a dashboards potential to inform decision-making. Seven features common to highly actionable dashboards were identified^2^.

To date, the experiences of dashboard developers (teams)—the actors responsible for a dashboard’s development—have predominately been captured through anecdotal descriptions, rather than structured evaluations of their development process. Recognizing the sustained importance of COVID-19 dashboards as a tool for pandemic reporting, opportunities for learning, exchanging experiences and co-designing recommendations for better preparedness in future public health crises is of critical importance.

### Aims

To describe the development of actionable COVID-19 dashboards from the perspective of their developers, including facilitating and hindering factors faced, and jointly identify lessons learned and recommendations for strengthening actionable public reporting in the context of a pandemic.

### Research questions

1. How can the development process of COVID-19 dashboards be described? Where this description includes decisions taken around the aim and audience, indicators selected, data sources used, links to policy measures, geographic breakdowns, population disaggregation, and use of visualizations^3^.
2. What facilitating and/or hindering factors were faced in the development of COVID-19 dashboards?

### Scope

To scope our investigation, we have put focus on *COVID-19 dashboards* that meet the following criteria: (i) reporting of key performance indicators related to COVID-19; (ii) use of some form of visualization (tables, maps, graphs); (iii) availability in an online, web-based format; (iv) reporting on COVID-19 in the scope of the 53 countries of the WHO European Region at the national level; and (v) developed by a government or appointed public authority with the responsibility to report pandemic-related information. See Annex 1 for the list of target dashboards identified.

### Approach

A qualitative study designed in two phases: (i) descriptive evaluation using semi-structured interviews with dashboard developers (individuals and/or their teams); and (ii) co-design of lessons learned and recommendations using workshop(s) with participants from phase (i).

### Study phases and timing

**Phase one: Semi-structured interviews with dashboard developers** (April and May 2021) At-distance interviews will be conducted by a member(s) of the study team in the preferred language of informants based on the working languages of the study team. Interviews are expected to last 60 minutes and will be structured around the two research questions to describe the dashboard’s development process and enabling and hindering factors faced. A detailed interview topic guide will be provided in advance.

**Phase two: Co-design workshop(s) with dashboard developers** (June 2021) Workshop(s) with participating dashboard developers will be convened in June 2021 to validate the analysis of findings resulting from phase one and to explore lessons learned for strengthening pandemic-related public reporting moving forward. The workshops are also an opportunity for developers to directly exchange with one another, making this phase both a learning and networking opportunity.

### Working languages

All study materials will be available in English and Russian. Interviews can be conducted in the preferred language of the interviewee, limited to the working languages of the study team: English, Russian, German, French, Bosnian, Croatian, Dutch, Italian, Montenegrin, Portuguese, Serbian, Slovenian, Spanish.

### Profile of key informants

Target key informants include individuals in senior strategic, operational, analytical or technical positions related to the development and running of COVID-19 dashboards in the WHO European Region (see Table enclosed). Informants ideally have involvement with the dashboard from its inception and have had oversight or influence over decisions related to its aim, content, data sources, display and dissemination. Interviews can be conducted jointly with one or more member of a dashboard’s team at the informant’s discretion.

### Dissemination and policy implications of results

It is the intention of the study team to submit the findings for peer-reviewed publication. Key informants will be noted in the acknowledgements unless requested otherwise. Dissemination is also foreseen to include a public webinar in fall 2021. Further details will follow. The study’s findings are expected to inform jointly developed lessons learned for actionable reporting using dashboards in the context of a public health crisis.

### Funding

This study is funded through the European Union’s Horizon 2020 research and innovation programme under grant agreement No. 765141.

### Study team and contact details

This study is developed by HealthPros in collaboration with the WHO Regional Office for Europe. The full study team includes HealthPros consortium members from the University of Amsterdam, OptiMedis AG, Corvinus University of Budapest, Scuola Superiore Sant’Anna, University of Oxford and University of Surrey. Contact details for the core study team are provided below.

### Are you interested to participate?

Are you involved in the strategic development and/or operations a public, web-based COVID-19 dashboard and interested to participate in this study? Alternatively, are you aware of the experts in your country that are? If so, please get in touch using this link to connect with the study team.

### Join our Launch Event 30th March 2021

#### COVID-19 public reporting from the perspective of dashboard developers

Tuesday March 30th | 12:00–13:00 (Copenhagen) | Zoom | English and Russian Please also join a public webinar where we will launch this study and present results from recent COVID-19 dashboard and health information system studies.

### Target COVID-19 national dashboards in WHO European Region Member States

The following lists COVID-19 dashboards identified in WHO European Region Member States that meet the study’s inclusion criteria of a public, web-based COVID-19 dashboard, reporting on the national level and developed by the government, ministry of health or a delegated public authority with the responsibility of publicly reporting COVID-19 dashboard. In some countries, more than one dashboard may meet these criteria. This list is not exhaustive nor definitive of the dashboards sought and can be adjusted based on the advice and expertise of country-specific informants.

| # | Country (dashboard link) | Organization (Developer) |
| --- | --- | --- |
| 1 | Albania | Ministry of Health and Social Protection |
| 2 | Andorra | Government of Andorra |
| 3 | Armenia | Ministry of Health of the Republic of Armenia |
| 4 | Austria | Bundesministerium für Soziales, Gesundheit, Pflege und Konsumentenschutz |
| 5 | Azerbaijan | Ministry of Health |
| 6 | Belarus | Ministry of Health of Belarus |
| 7 | Belgium | Sciensano |
| 8 | Bosnia and Herzegovina | Government of the Federation of Bosnia and Herzegovina, Bosnia and Herzegovina |
| 9 | Bulgaria | Ministry of Health |
| 10 | Croatia | Government of the Republic of Croatia |
| 11 | Cyprus | *No applicable dashboard identified* |
| 12 | Czechia | Ministry of Health |
| 13 | Denmark | State Serum Institute |
| 14 | Estonia | Social Ministry |
| 15 | Finland | Finnish Institute for Health and Welfare (THL) |
|  | France | Government of France |
| 16 | France | Santé Publique France (Public Health France) |
| 17 | Georgia | *No applicable dashboard identified* |
| 18 | Germany | Robert Koch-Institut |
| 19 | Greece | *No applicable dashboard identified* |
| 20 | Hungary | Government of Hungary |
| 21 | Iceland | Directorate of Health |
| 22 | Ireland | Government of Ireland |
| 23 | Israel | Ministry of Health |
| 24 | Italy | Department of Civil Protection |
|  | Italy | National Agency for Regional Health Services (AGENA S) |
| 25 | Kazakhstan | Ministry of Health |
|  | Kazakhstan | National Center for Public Health |
| 26 | Kyrgyzstan | Ministry of Health of Kyrgyz Republic |
| 27 | Latvia | Center for Disease Prevention and Control |
| 28 | Lithuania | State Enterprise Centre of Registers (Registru Centras) |
| 29 | Luxembourg | The Luxembourg Government |
| 30 | Malta | Ministry of Health of Malta |
| 31 | Monaco | *No applicable dashboard identified* |
| 32 | Montenegro | Public Health Institute of Montenegro |
| 33 | Netherlands | National Institute for Public Health and the Environment (RIVM) |
|  | Netherlands | Rijksoverheid |
| 34 | North Macedonia | *No applicable dashboard identified* |
| 35 | Norway | Norwegian Directorate of Health |
|  | Norway | Norwegian Institute of Public Health |
| 36 | Poland | Government of Poland |
| 37 | Portugal | Directorate-General of Health |
| 38 | Republic of Moldova | Ministry of Health, Labour and Social Protection |
| 39 | Romania | National Institute for Public Health |
|  | Romania | Romanian Ministry of Interior |
| 40 | Russian Federation | Ministry of Health of Russian Federation |
| 41 | San Marino | Institute for Social Security |
| 42 | Serbia | Serbian Ministry of Health and the Institute of Public Health of Serbia |
| 43 | Slovakia | Ministry of Health of the Slovak Republic |
| 44 | Slovenia | National Institute of Public Health |
|  | Slovenia | Government Communication Office |
| 45 | Spain | Ministry of Health |
|  | Spain | National Center of Epidemiology |
| 46 | Sweden | Swedish Intensive Care Registry (SIR) |
|  | Sweden | Public Health Agency of Sweden (Folkhälsomyndigheten) |
| 47 | Switzerland | Bundesamt für Gesundheit BAG |
| 48 | Tajikistan | Ministry of Health and Social Protection of the Republic of Tajikistan |
| 49 | Turkey | Ministry of Health of Turkey |
| 50 | Turkmenistan | *No applicable dashboard identified* |
| 51 | Ukraine | Ministry of Health |
| 52 | United Kingdom | Public Health England |
| 53 | Uzbekistan | Ministry of Health |

## Appendix 2: Brief interview topic guide

### 1. Development process and team

- Briefly, please describe the development process of your dashboard, specifically key milestones from its inception to its launch as well as any main changes that have taken place over the course of 2020 to present-day.
- Describe the members of the team that work on the dashboard on a regular basis, specifically, the competencies (profile) of these individuals, key experts, organizations or stakeholders collaborated with and any significant changes to the team over time.

### 2. Description of the dashboard’s key features^1^

- **Purpose and users**. Was the purpose of the dashboard defined from the start? Who was considered the intended user? Have any measures to track the use of the dashboard been taken?
- **Information (indicators)**. What was the process for deciding which indicators to report on? Who was involved in that process? How was the ordering and clustering of indicators decided on?
- **Data sources and methods**. How were the sources of data selected? What permissions to gain access were involved? What data would you have liked to have had access to?
- **Reporting data over time**. To what extent were infection control policy measures (e.g., mandatory use of masks) reported on the dashboard to show their effect over time? Why or why not was this done?
- **Geographic breakdowns**. How has the geographic breakdown of data (e.g., national, regional, municipal, post code-level) changed over time? Did more granular data become available? In your opinion, what breakdowns are missed and why?
- **Population breakdowns**. What population breakdowns were applied (e.g., sex, age, ethnicity)? How did these change over time? In your opinion, what breakdowns are missed? Why?
- **Visualizations and explanations**. How were displays decided upon (e.g., charts, tables, graphs)? How have you used visual cues and explanations on the data and trends to improve readability? Have you tested the readability/user experience of the dashboard?

### 3. Summary of key barriers and enabling factors and lessons learned

- In your opinion, what has been the most advantageous factor supporting the development and running of the dashboard? What was the most challenging factor and/or greatest barrier faced in the development and running of the dashboard?
- With the benefit of hindsight, what would you do differently? What advice would you offer to other countries to best prepare for public reporting in the context of a public health crisis?
- What are your plans for the dashboard for the remainder of 2021?

## Appendix 3: Sample details

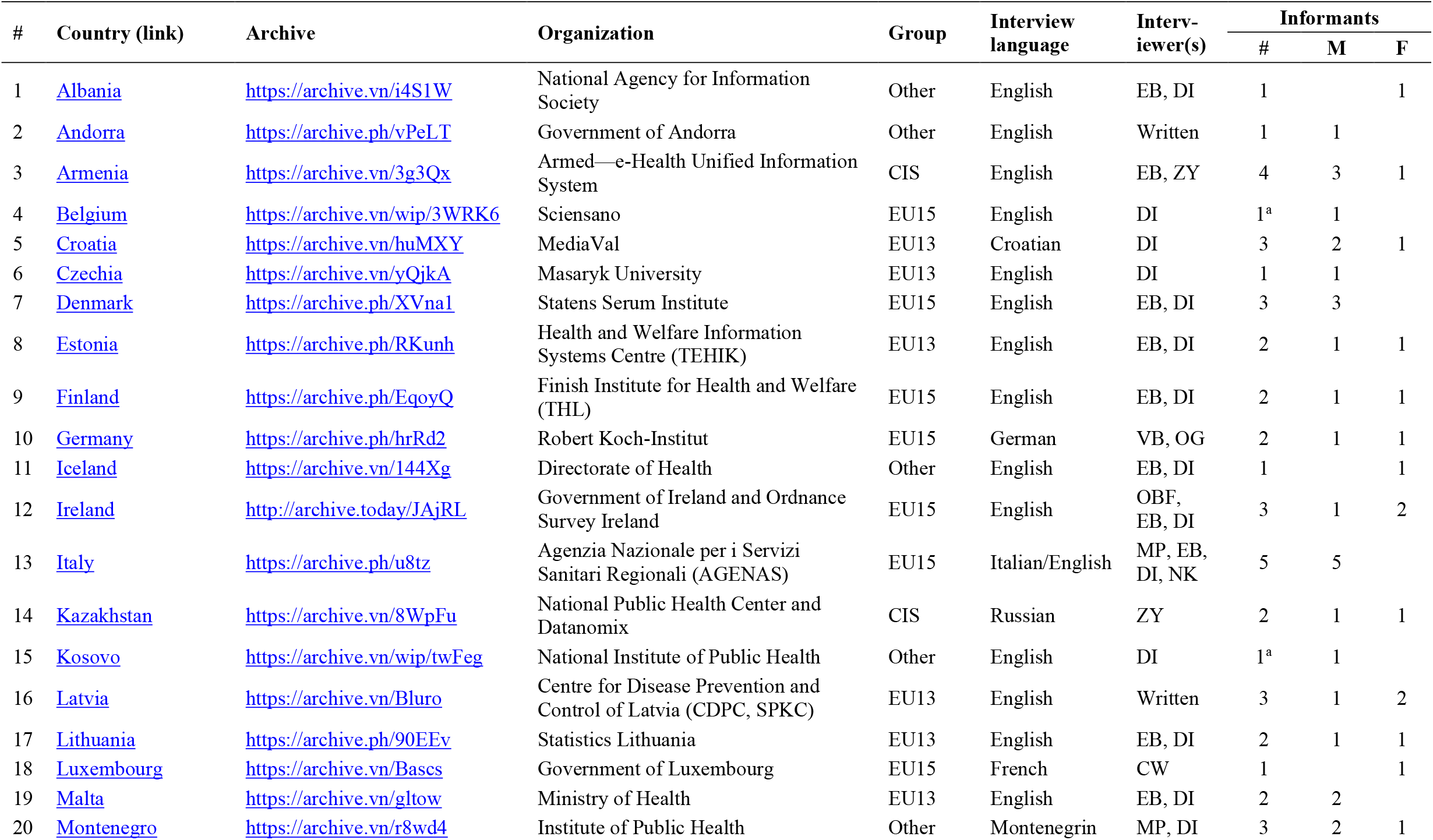
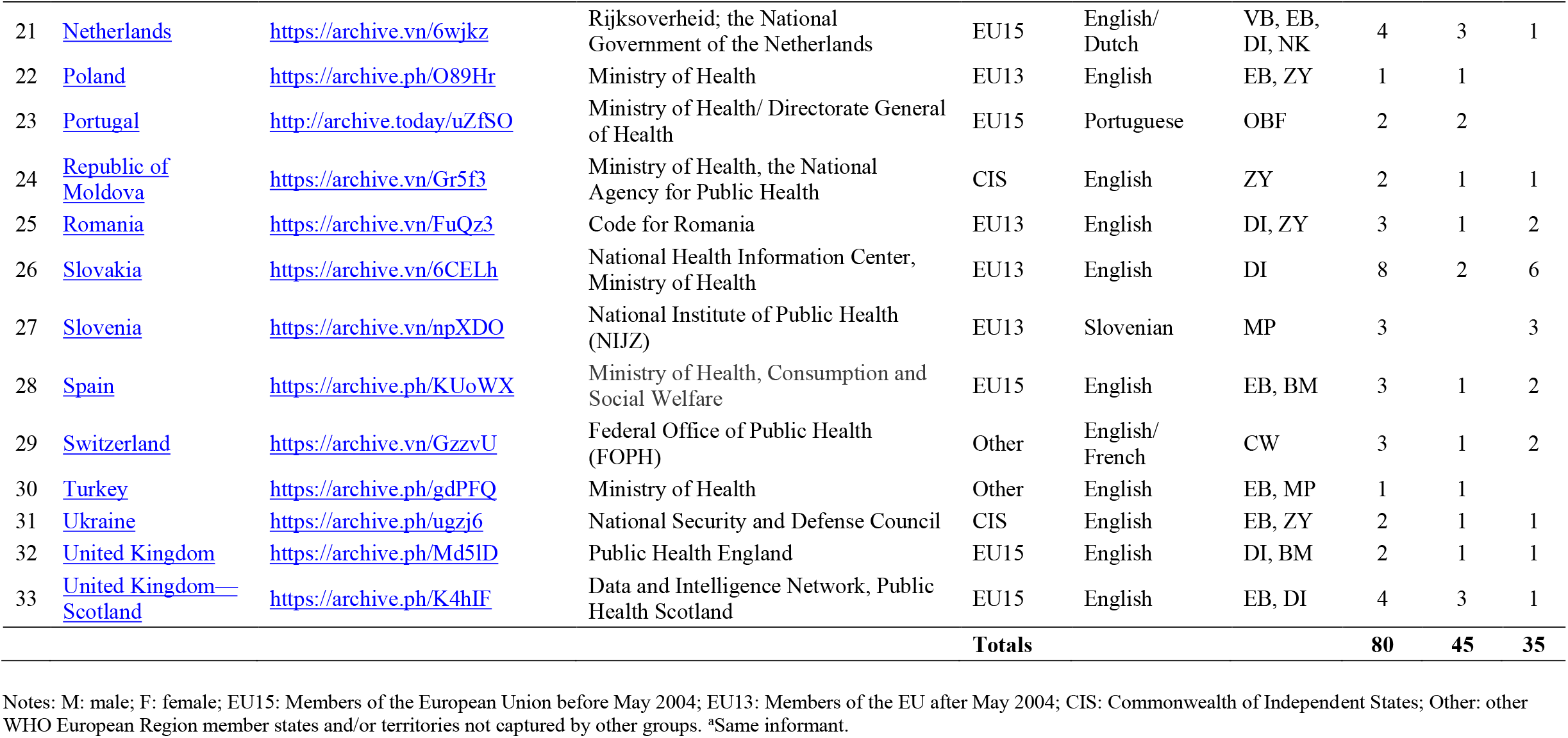

## Footnotes

1 Marie Skłodowska-Curie Innovative Training Network for Healthcare Performance Intelligence Professionals running 2018-2022. The network sets out to train a cohort of 14 HealthPros Fellows. The network’s consortium spans partners in Canada, Denmark, Germany, Hungary, Italy, the Netherlands, and United Kingdom. For more information on the HealthPros network, visit: https://www.healthpros-h2020.eu/.

2 Ivanković D, Barbazza E, Bos V, Fernandes OB, Gilmore KJ, Jansen T, Kara P, Larrain N, Lu S, Torres BM, Mulyanto J, Poldrugovac M, Rotar A, Wang S, Willmington C, Yang Y, Yelgezekova Z, Allin S, Klazinga NS, Kringos DS. Features Constituting Actionable COVID-19 Dashboards: Descriptive assessment and expert appraisal of 158 public, web-based COVID-19 dashboards. JMIR; 2021. https://www.jmir.org/2021/2/e25682/.

3 This description reflects the seven features of highly actionable dashboards: (1) know the audience and their information needs; (2) manage the type, volume and flow of displayed information; (3) report data sources and methods clearly; (4) link time trends to policy decisions; (5) provide data ‘close to home’; (6) breakdown the population into relevant sub-groups; and (7) use story-telling and visual cues. Refer to the article for the description in full: https://www.jmir.org/2021/2/e25682/.

1 The features highlighted for discussion draw from our previous study on a global sample of COVID-19 dashboards. Find the full article here: https://pubmed.ncbi.nlm.nih.gov/33577467/.

